# “The System is Overwhelming and Frustrating and Discouraging”: Stress and Social Support for Affected Family Carers in a Toxic Drug Crisis

**DOI:** 10.1101/2025.10.20.25336204

**Authors:** Saranee Fernando, J. Hawkins, A. Salmon, M. Kniseley, S. Esau, M. Sikora, C. Long, J. Robson, D. Snyder

## Abstract

**Background:** Family members and other affected carers are essential supports for the wellbeing of people who use drugs who are at risk of an overdose. This study explores the perspectives of affected family carers of loved ones at risk of an overdose or who experienced a past fatal overdose in the Fraser East region of British Columbia (BC).

**Methods:** In this qualitative study, collaborative thematic analysis of 22 interviews was conducted examining the experiences of family carers of loved ones at risk of death due to drug toxicity. This study was guided by community-based participatory action research (CB-PAR) principles by involving peer researchers at all research stages.

**Results:** Family carers experienced tremendous stress while supporting their at-risk loved ones, particularly relating to stress caused by the toxic drug supply. Family carers also encountered substantial stress when navigating healthcare services, engaging with substance use services and finding support in the Fraser East region of BC. Their experiences were marked by traumatic episodes and severe instability, leading to crises such as damaged relationships and overdose incidents of loved ones.

**Conclusion:** The findings of this study offer unique insights into the distinct stressors and support experiences of family carers, with particular focus on the toxic drug crisis and the fragmented nature of both informal social networks and formal support systems. The stress-strain-coping-support (SSCS) model is discussed as a useful framework for describing the needs and experiences of family carers of those at risk of overdose within the context of the toxic drug crisis.

## Introduction

In 2016, the province of British Columbia (BC) declared a public health emergency in response to a rapid increase in the number of people dying due to poisoning of the unregulated drug market with fentanyl analogues, which has become known as the “toxic drug crisis”. Since that time, more than 16,000 people have died due to drug toxicity in the province (BC Coroner’s Service, 2025). An important driver of these continuing deaths is the lack of evidence-based interventions to address the needs of people who use drugs (PWUD) alone in private residences in rural and semi-urban communities (BC Coroner’s Service, 2024; Fernando et al., 2022; Papamihali et al., 2020).

Our initial community-based participatory research (CB-PAR) study based in the Fraser East region of BC revealed that as their drug use increased, many PWUD became increasingly isolated, and their relationships with family members and close friends deteriorated (Fernando et al., 2022). Family carers also report experiencing extreme stress when supporting their loved ones struggling with substance use and emphasize that they require support in their own right (Copello et al., 2010; Denomme & Benhanoh, 2017; Hal, 2012; Hornberger & Smith, 2011). Compounding the issue, family carers often experience internal dissonance regarding relational negotiations with their loved ones at risk of overdose (Hawkins et al., 2025). Despite their need for support, however, family members affected by their loved one’s substance use often retreat from their community due to stigma, guilt, and shame (Coemans et al., 2015).

Since 2018, our team has been engaged in CB-PAR research aimed at identifying effective responses to the toxic drug crisis in the Fraser East region of British Columbia. Our focus has been on reducing deaths among people who use drugs (PWUD), particularly those resulting from unwitnessed drug poisonings. Recognizing the vital role that close family members and other loved ones can play in preventing such deaths, especially when drug use occurs in isolation (Fernando et al., 2022), the present study explores the experiences of family carers of PWUD in Fraser East whose loved ones have experienced unwitnessed drug poisonings.

This study focuses specifically on stress and support because family members often occupy a dual role of caregivers and emotionally impacted witnesses to the ongoing crisis, requiring support in their own right (Orford et al., 2010). While family carers may serve as first responders in their loved one’s overdose events or provide critical ongoing support, their own mental health and coping needs are often unaddressed. Yet supportive families and other loved ones have been recognized as essential supports for the wellbeing of PWUD (Fernando et al., 2022; Mathias et al., 2024).

## Materials and Methods

Guided by community-based participatory research (CB-PAR) principles (Closson et al., 2016; Wilson, 2019), our research team is composed of people with a history of drug use, family carers of PWUD, public health physicians, representatives of municipal governments and the regional health authority, representatives of non-profit organizations, community health specialists, and academics as co-researchers in the rural and semi-urban Fraser East region.

The study utilized purposive and respondent-driven sampling methods to identify and engage potential participants. Referrals were facilitated by community partners through word-of-mouth, and connections were established via social networks by peer research associates (PRAs) and research team members. Participants who referred up to two individuals who met the eligibility criteria received a $10 incentive per referral. This referral cap ensured sample diversity by limiting the number of participants recruited through any single respondent. Eligible participants were individuals aged 19 and over who were substance-affected family carers, identified as those currently supporting a loved one or had lost a loved one in the rural and semi-urban Fraser East region since the onset of the public health emergency in April 2016.

Interviews were conducted by PRAs at a safe location chosen by the participant within their community or over the phone, and each participant received a $50 cash honorarium for their participation. Interviews were conducted in various settings, including participants’ homes and public spaces such as coffee shops and community centers. Prior to each interview, the interviewers guided participants through an informed consent process that emphasized privacy and confidentiality. During this process, self-reported information was collected on participant demographics and relationship to the person using at risk of overdose. Interview questions delved into the relationship between the family carer and their loved one at risk of OD, experiences of connection and isolation of the loved one, system interactions, the overdose event(s), and supports accessed by the family carer.

In total, semi-structured interviews (n=22) were conducted by the team members with lived experience, supported by a research assistant based in the Fraser East. Interviews were conducted between August 2020 and August 2021, audio recorded using an encrypted device, and stored on a secure server. Transcripts were generated using de-identified audio files marked by participant pseudonyms. As interviews occurred during the COVID-19 pandemic, public health guidelines were adhered to at all stages of data collection. Ethics approval was granted by the UBC Providence Health Care Research Ethics Board (H18-02881).

This study adopted a social constructivist lens, recognizing that knowledge is co-constructed between the researcher and participants. Data analysis was oriented towards a constructivist epistemology, with an experiential orientation to data analysis that emphasized meaning expressed by participants. Reflective Thematic Analysis was conducted collaboratively between two researchers (J.H, S.F.). Themes were identified deductively and inductively with analysis aided by NVivo software. A master codebook was created by the primary coder (J.H), who created and validated the master codebook with the input of the research team. Data validation occurred through a second round of coding (S.F). The codebook was discussed between coders and resolutions were made collaboratively. The process also involved peer-led member checking through data discussions, leveraging the embedded nature of peer researchers in the community. To illustrate, analyses were presented to the larger team, including PRAs, who had reviewed deidentified interview transcripts. Sense-making meetings were held with the research team to discuss and interpret the emerging trends from the data. This collaborative process allowed for further elaboration and refinement of the findings.

## Results

The total of 22 participants included six parents, three siblings, one cousin, four intimate partners/spouses, and eight friends of people who use drugs (PWUD). Fifteen participants identified as female and seven as male; none identified as another gender. Fifteen had lived experience with substance use, and seven participants were involved in paid frontline work in substance use services. Six participants’ loved ones were deceased, and 15 loved ones continued to be at risk at the time of the interview. One participant experienced both a loss of a spouse to overdose and a child who continued to be at risk.

Overall, family carers bore a significant weight of stress while tending to their at-risk loved ones, particularly relating to stress caused by the toxic drug supply and the systems currently available for addressing the crisis. Family carers experienced substantial stress burdens when navigating healthcare services, seeking social support, and engaging with substance use services (i.e. detoxification centres, substance use management services) within the Fraser East region of BC. Their life trajectories included moments of trauma and extreme chaos, precipitating crises such as strained relationships and overdose events by their loved ones.

### Stress and the toxic drug supply

“I just know a lot of people are overdosing lately, and that most of it is fentanyl.” – Dylan Participant narratives described experiences of worry, relational deterioration and loss uniquely situated within the context of highly toxic drugs. Throughout participant narratives, heightened anxiety was pervasive, stemming from the awareness of the toxic drug supply. This concern was exacerbated by the knowledge of fentanyl’s devastating effects within the broader community, or even within their own social circles and families. Numerous individuals expressed their apprehension regarding the potentially lethal repercussions of consuming fentanyl-contaminated substances, reflected in this comment from a friend (Audrey): “Things have changed so much over the years and it just seems like it’s getting worse and worse and it’s scary. It’s just scary.” This anxiousness was a common undercurrent across accounts, intensifying the stress felt by family carers, as the event of overdose was more likely to occur than before. As described by a man who lost a family member to overdose (Greg): “And chances are at this day and age with the drugs that are hitting the street, they’re going to do carfentanil or fentanyl […] and you’re going to have another carcass that’s sitting in a fucking morgue.”

Family carers also navigated a profound sense of loss stemming from personality changes that they believe to be unique to fentanyl use, and they spoke about the toll of supporting a loved one with these stark and intense behavioural shifts. One participant remarked on the distinctive nature of fentanyl compared to other illicit drugs:

> Fentanyl, it’s a different thing. It’s not heroin. Not even remotely. People don’t act the same. They have, like I can, I’ll see a person with, you know, morals and standards and everything and then do some fentanyl. And that all goes right out the window. […] Like fentanyl does something to people’s noodles. Even if they do quit or if they do get on a substitute, they’re just not the same. - Evelyn

### Stress in the context of acute crisis

“I have a Facebook account full of dead people from overdoses. My son overdosing like 20 times in the course of two years.” - Maddison

The overdose crisis presents unprecedented instances of loss bearing directly on experiences of overdose events. Family carers grappled with the aftermath of witnessing multiple overdoses, finding their loved one deceased or having to resuscitate their loved one from near death.

Overdose events often led to a chaotic cascade of traumatic events, and in many cases, an ambulance was called to the scenes which were common crisis points for carers, who were desperate to help their loved ones survive an overdose event:

> I mean, there’s been numerous instances where you find people essentially almost dead, and you call 911 and sit there and watch somebody do chest compressions on them, really chest compressions on them, while you don’t know if they’re going to live or die. That fucks with a person’s head…-Matt

When describing overdose events, family carers highlighted the rippling impact of a loved one’s risky substance use on larger family systems, as indicated by a spouse of an individual who died of an overdose (Jess): “So yeah, so when you, you know, you think it just affects the drug addict, it doesn’t. You know, there’s a son, a grandson, daughter-in-law, son-in-law, his mum, brother, sister; there’s a whole group of family affected, just because of one person.”

In numerous cases, family carers expressed mental fatigue and numbness towards a crisis of increasing frequency of overdoses in the community, particularly if they were also working or volunteering in the context of the crisis, as mentioned by a mother of a son who experienced multiple non-fatal overdoses (Madison): “It used to be like when somebody would die, it was emotionally traumatizing for me. And because it’s happened so much over the last four years, I’d say that—and this is a really shitty thing to say—but it’s almost expected.”

### Challenges with substance use systems

Family carers, notably parents and spouses, recounted numerous negative encounters with healthcare and social services, resulting in feelings of exhaustion, frustration, emotional distress, and disillusionment in the system. The scarcity of beds at treatment facilities or withdrawal management services was highlighted as a particularly vexing issue for both family carers and PWUD, particularly by family carers who also worked in social services (Madison): “What should happen is we need more detox centres. We need more beds. We need more treatment centres. We need it to be easier to get into detox. Like to get into detox, you’ve got to phone them every day for two or three weeks.”

Grappling with the complexities of accessing care and fulfilling the necessary requirements for loved ones facing overdose risks compounded frustrations and eroded trust in the system. This sentiment was articulated by a mother whose son was at risk of overdose (Alice): “I find the system [is] overwhelming and frustrating and discouraging. They’re not, the systems aren’t designed around people who use substances, they’re, I don’t know who they’re designed for, but it’s not for their supposed clients.”

Family carers expressed a profound sense of frustration upon observing interruptions in care or encountering refusals of service:

> It has not helped when I have to start from scratch and start reaching out to get help and then be put on hold and then redirected, and then finally get a—And while the trauma and everything’s red hot finally get someone while the trauma’s done most of its damage. And then I get to someone who – I’m just one more in a line-up in their day and we don’t even know each other and he’s going to try and make an assessment based on one hour. […] So, I find for me if I can get any help any other way I’ll avoid that path. - Doug

### Informal social supports to alleviate stress burdens

“I wouldn’t still be alive right now, I believe, if it wasn’t for my mother’s continued unconditional love and support.” - Patrick

Informal supports were often established through relationships with family members or friends with similar lived experience. These networks proved invaluable in providing support in various capacities amidst multiple confounding challenges. Peers with shared lived experiences provided a safe space for family carers to receive non-judgmental support, as mentioned by a parent of a son at risk of overdose (Anna): “I’ve gone actually to other women in my life who have a certain amount of experience and our walk and talks have been very helpful.”

In contrast, often family carers’ primary social networks did not offer them as much support. This is reflected in the account of a sister of a woman at risk of overdose (Callie): “My partner’s not the person I go to for support unfortunately. It’s not the way things should be, but that – he’s the last person I would go to.”

Some individuals sought solace through engagement with spirituality and faith-based leaders, as indicated by the following account from a friend whose loved one died (Audrey): “The pastor is awesome and stuff and they just remind me that I am loved and you know, Jesus is there for me and I truly believe that he is there for all of us.” Furthermore, having shared faith in the community was valued as the act of praying together was important during times of crisis, as recounted by a close friend (Doug): “[The overdose] was discovered early in the morning and it was friends, friends close by had become aware of it almost immediately and, you know, a lot of us share a common faith so prayer becomes a very important matter.” Some accounts highlighted that mixed spiritual approaches were used to help mitigate stress and mental health burdens, as illustrated by a friend of a woman at risk of an overdose (Kendra): “I think just between – like I practice Buddhist concepts which is why I think that works, and then the AA (*Alcoholics Anonymous*) spiritual way of life.”

### Challenges with formal supports

More than half of the participants described accessing formal supports within the community such as community services, support groups, and counselling. Several participants with lived experience of substance use were on methadone and accessed other formal supports for their own wellness such as supportive housing programs or 12-step programs. Professionals did not always seem prepared to encounter the complexity of the family carer burden. One woman whose husband died of drug overdose spoke of her own confusion navigating the complexities of their relationship that did not seem mitigated by the counsellors she accessed:

> I certainly went to a number of counsellors, most of them were doing that approach, sort of took that standpoint where you have to let them hit rock bottom which, I don’t know, I didn’t do that for many, many years and maybe that – maybe just – honestly when he finally did go to rehab it was when we left. So, I don’t know. I don’t know what way is better. I’m not sure. I still – I have no idea. But then maybe in the end he didn’t tell me because he was worried that we would leave again. So, I don’t know. - Melissa

At the same time, this wife and mother also persevered in accessing counselling to ameliorate the adverse effects of trauma, exhibiting resiliency in the face of adversity:

> And trying to deal with everything that’s coming your way […] I’ve been to a number of counsellors and – since – probably 2012-ish. And yeah, I think most people sort of look at me and they’re sort of saying, how did you survive what you did? Like, the stress level that you had in your daily life was immense. And yeah, so how did you survive that? - Melissa

## Discussion

This study highlighted significant contextual factors that impact family carer stress, particularly during the toxic drug crisis and its associated strain on support systems. The unpredictable and increasingly lethal nature of the unregulated drug market has created a pervasive sense of fear and uncertainty for family carers of people who use drugs. For family carers, this means living in constant vigilance and intensified chronic stress, never knowing if their loved one will survive each use, especially when they are using alone. The risk of fatal overdose permeates everyday life with crisis management, forcing carers to make agonizing decisions about their level of involvement with their loved one. The ongoing realities of the overdose crisis also present an additional layer of tragedy and complexity. Participant accounts were dense with detailed descriptions of traumatic events that were often not helpfully or comprehensively addressed by formal supports, while often even exacerbated by them. Ultimately, the toxic drug crisis significantly amplifies affected family member stress, often leaving carers feeling powerless, isolated, and insufficiently supported by social networks and formal support systems.

### Evidence-based approaches – the SSCS model

The findings from this study point toward urgent needs for evidence-based approaches that recognize the strengths and needs of family carers. In many Canadian jurisdictions, supports for family members caring for a loved one with substance use problems are limited, fragmented, or nonexistent (Denning, 2010; O’Connor, 2021). In addition, the paradigms in which many supports for family carers are grounded have been identified as problematic and disenfranchising, whereby families are conceptualized as dysfunctional contributors to their loved one’s problematic substance use (Devaney, 2017; Orford et al., 2013). For example, codependency theory (Bacon et al., 2020), which influences popular discourse and normative understandings of substance use within families (Bacon et al., 2020; Calderwood & Rajesparam, 2014; Denning, 2010; Rotunda & Doman, 2001), pathologizes family members supporting loved ones by reproducing narratives of emphasizing that their loved one’s recovery can only be achieved through “tough love” and “hitting rock bottom” (Copello et al., 2010; Hawkins et al., 2025; Rotunda & Doman, 2001). At the same time, codependency theory rejects family members’ efforts to intervene in ways that reduce harms for their loved ones as “enabling” their substance use. Co-dependency theory is widely adopted in practice despite a lack of standardized definitions and assessment metrics, the presence of over-abundant diagnostic criteria falling under “co-dependency”, the emergence of questions surrounding its therapeutic utility, and feminist critique (Bacon et al., 2020; Calderwood & Rajesparam, 2014; Dear & Roberts, 2005).

In contrast to approaches that pathologize family carers and their efforts to support PWUD, Orford et al’s stress-strain-coping-support (SSCS) model offers an alternative way to understand and address family carers’ stress. The SSCS model proposes that caring for a close relative with a substance use problem is a chronic, stressful experience that can result in family carer strain, manifesting in carers’ ill-health (Orford, Velleman, et al., 2010). This stress and strain can be mitigated by the presence of effective supports and resources which improve a family carer’s ability to cope and, in turn, their ability to care well for their loved ones experiencing substance use problems. In the SSCS paradigm, coping can be seen as adaptive or maladaptive in terms of its outcome, but is not pathologized as an expression or response (McCann & Lubman, 2018).

In the SSCS model, an absence of support or coping capacity is understood to fundamentally undermine a family carer’s wellbeing, and thus their capacity to continue supporting their loved one; however, in contrast to many existing models, the SSCS model centers the importance of acknowledging the stress and strain experienced by family carers who support PWUD without requiring direct participation of the family member they are supporting (Copello et al., 2009; Kourgiantakis & Ashcroft, 2018). This decreases barriers to accessing support for family carers, who are already under-supported in the context of the toxic drug crisis (Mathias et al., 2025). By increasing non-pathologizing support available to family carers, the SSCS proposes that family carers will experience enhanced resiliency in supporting their loved one (Denomme & Benhanoh, 2017).

### Finding support during a toxic drug crisis

Social support has been identified as a key factor affecting family carer wellbeing and coping (Hellum et al., 2022). The SSCS model posits that social support can be informal (e.g., offered by family members, friends, and partners) or formal (e.g., offered by professionals such as physicians, social workers, or agencies) and received as emotional, material or informational (Orford, Copello, et al., 2010). Participants illustrated efforts in seeking and utilizing both informal and formal support systems to alleviate the emotional burdens of stress. Material support was not as prominently sought or received by participants in this study, but many participants spoke of increasing their knowledge of addiction as a key alleviating factor.

In many cases, family carers’ primary social networks were not perceived as offering them support, which can be a common experience (Orford, Velleman, et al., 2010). However, informal supports involving peers with lived experience of substance use were highlighted as particularly helpful.

Consistent with previous research, our findings suggest that families can act as a mediating influence in the lives of PWUD, supporting them through their substance use (Fernando et al., 2022). However, this role is often complicated by systemic barriers to formal supports such as timely and accessible care services, a safer drug supply, detoxification beds, intensive substance use treatment, and professionally-supported peer social support networks. In our study, the majority of participant interactions with substance use service systems were reported to have brought further stress. In fact, while the SSCS model does reference the stress that systems can cause (Orford, Velleman, et al., 2010), the many narratives in this study that speak to adverse experiences navigating substance use service systems constituted its own category of stress rather than support. Nevertheless, formal interventions can provide key support by preventing carers and their loved ones from reaching crises such as unwitnessed drug poisonings (Fernando et al., 2022) and addressing specific and acute crisis points as they arise, particularly through providing evidence-based information (i.e. naloxone training) (Williams et al., 2014) and facilitating peer-based social support.

There is a paucity of research centering family carers in the context of the toxic drug crisis. This work brings new insights into factors of stress and support in the SSCS model, tracing the unique emotional, relational, and systemic burdens of caregiving during a public health emergency. Understanding family carer experiences of stress and support is essential for informing interventions and policies that extend care beyond the individual who uses drugs to include their immediate and impacted social networks. Based on these findings, interventions should incorporate social supports, such as peer navigators, that can aid family carers in navigating the challenges of supporting a loved one at risk of toxic drug poisoning. Additionally, peer-based social supports, such as peer support groups for families affected by the toxic drug crisis, can create a safe emotional space where individuals feel comfortable sharing their struggles and heightened anxiety surrounding the toxic drug supply. Spirituality and faith communities can also offer family carers solace under stressful circumstances (Orford et al, 2013). More efforts should be made to engage faith communities (Banks et al., 2023) to enhance their ability to address the toxic drug supply crisis, such as through educational programs, naloxone trainings, or dialogue sessions.

By enhancing access to peer-based social support and relevant information, interventions informed by the SSCS model can bolster the resilience of family carers as they navigate the fraught challenges of supporting their loved ones.

## Acknowledgements

We are thankful for the continued support of our research team and networks supporting this work, including community advisors, peer research associates, and community members residing in the Fraser East region of BC.

## Declaration of Interest Statement

The authors report there are no competing interests to declare.

## Data availability

Data was collected and managed in concordance with the University of British Columbia Research and Providence Health Care Ethics Boards, which do not permit access to this data. Requests to access data for secondary analysis may be submitted to the corresponding author upon reasonable request.

## Funding

This work was funded by the Vancouver Foundation under the Participatory Action Convene Grant F18-03489, the Canadian Institute of Health Research under the Catalyst Grant F19-04007, and the Canadian Institute of Health Research under the Project Grant F22-01474.

## Notes

### Competing Interest Statement

The authors have declared no competing interest.

### Funding Statement

This work was supported by the Vancouver Foundation under the Participatory Action Convene Grant F18-03489, the Canadian Institute of Health Research under the Catalyst Grant F19-04007, and the Canadian Institute of Health Research under the Project Grant F22-01474.

### Author Declarations

The University of British Columbia Providence Health Care Research Ethics Board of the University of British Columbia and Providence Health Care gave ethical approval for this work.(H18-02881)

### Summary of Updates

Discussion of the SSCS model was moved from the introduction to the discussion.

## References

Bacon, I., McKay, E., Reynolds, F., & McIntyre, A. (2020). The Lived Experience of Codependency: an Interpretative Phenomenological Analysis. International Journal of Mental Health and Addiction, 18(3), 754–771. 10.1007/s11469-018-9983-8

Banks, D. E., Brown, K., & Saraiya, T. C. (2023). “Culturally Responsive” Substance Use Treatment: Contemporary Definitions and Approaches for Minoritized Racial/Ethnic Groups. Current Addiction Reports, 10(3), 422–431. 10.1007/s40429-023-00489-0

BC Coroner’s Service. (n.d.). Illicit Drug Toxicity Deaths in BC (January 1, 2011-December 31, 2021). https://www2.gov.bc.ca/assets/gov/birth-adoption-death-marriage-and-divorce/deaths/coroners-service/statistical/illicit-drug.pdf

Calderwood, K. A., & Rajesparam, A. (2014). A Critique of the Codependency Concept considering the Best Interests of the Child. Families in Society: The Journal of Contemporary Social Services, 95(3), 171–178. 10.1606/1044-3894.2014.95.22

Closson, K., McNeil, R., McDougall, P., Fernando, S., Collins, A. B., Baltzer Turje, R., Howard, T., & Parashar, S. (2016). Meaningful engagement of people living with HIV who use drugs: methodology for the design of a Peer Research Associate (PRA) hiring model. Harm Reduction Journal, 13(1), 26. 10.1186/s12954-016-0116-z

Coemans, S., Wang, Q., Leysen, J., & Hannes, K. (2015). The use of arts-based methods in community-based research with vulnerable populations: Protocol for a scoping review. International Journal of Educational Research, 71, 33–39. 10.1016/j.ijer.2015.02.008

Copello, A., Orford, J., Velleman, R., Templeton, L., & Krishnan, M. (2000). Methods for reducing alcohol and drug related family harm in non-specialist settings. Journal of Mental Health, 9(3), 329–343. 10.1080/JMH.9.3.329.343

Copello, A., Templeton, L., Orford, J., & Velleman, R. (2010). The 5-Step Method: Evidence of gains for affected family members. Drugs: Education, Prevention and Policy, 17(Sup1), 100–112. 10.3109/09687637.2010.514234

Copello, A., Templeton, L., Orford, J., Velleman, R., Patel, A., Moore, L., MacLeod, J., & Godfrey, C. (2009). The relative efficacy of two levels of a primary care intervention for family members affected by the addiction problem of a close relative: a randomized trial. Addiction, 104(1), 49–58. 10.1111/j.1360-0443.2008.02417.x

Dear, G. E., & Roberts, C. M. (2005). Validation of the Holyoake Codependency Index. The Journal of Psychology, 139(4), 293–314. 10.3200/JRLP.139.4.293-314

Denning, P. (2010). Harm reduction therapy with families and friends of people with drug problems. Journal of Clinical Psychology, 66(2), 164–174. 10.1002/jclp.20671

Denomme, W. J., & Benhanoh, O. (2017). Helping concerned family members of individuals with substance use and concurrent disorders: An evaluation of a family member-oriented treatment program. Journal of Substance Abuse Treatment, 79, 34–45. 10.1016/j.jsat.2017.05.012

Devaney, E. (2017). The emergence of the affected adult family member in drug policy discourse: A Foucauldian perspective. Drugs: Education, Prevention and Policy, 24(4), 359–367. 10.1080/09687637.2017.1340433

Fernando, S., Hawkins, J., Kniseley, M., Sikora, M., Robson, J., Snyder, D., Battle, C., & Salmon, A. (2022). The Overdose Crisis and Using Alone: Perspectives of People who use Drugs in Rural and Semi-Urban areas of British Columbia. Substance Use and Misuse.

Hal, S. H. G. van. (2012). Experiences of parents of substance-abusing young people attending support groups. Archives of Public Health, 70(11).

Hawkins, J; Salmon, A; Fernando, S; Battle, C.; Esau, S.; Snyder, D; Sikora, M. (2025). ‘I don’t know what we should have done differently’: A qualitative study on the dilemmas of ‘tough love’ and toxic drugs in British Columbia, Canada. Drugs: Education, Prevention and Policy, 1–10. 10.1080/09687637.2025.2493140

Hellum, R., Bilberg, R., & Nielsen, A. S. (2022). “He is lovely and awful”: The challenges of being close to an individual with alcohol problems. Nordic Studies on Alcohol and Drugs, 39(1), 89–104. 10.1177/14550725211044861

Hornberger, S., & Smith, S. L. (2011). Family involvement in adolescent substance abuse treatment and recovery: What do we know? What lies ahead? Children and Youth Services Review, 33, S70–S76. 10.1016/j.childyouth.2011.06.016

Kourgiantakis, T.; Ashcroft, R. (2018). Family-focused practices in addictions: A scoping review protocol. BMJ Open, 8(1). 10.1136/bmjopen-2017-019433

Lee, K.M.T.; Manning, V.; Teoh, H.C.; Winslow, M.; Lee, A.; Subramaniam, M.; Guo, S.; Wong, K.E. (n.d.). Stress-coping morbidity among family members of addiction patients in Singapore. Drug and Alcohol Review, 30(4), 441–447. 10.1111/j.1465-3362.2011.00301.x

Mathias, H.; Auger, S., Schulz, P.; Hyshka, E. (2025). Including families in a response to the unregulated toxic drug crisis: A call to action. 60(3), 452–456. 10.1080/10826084.2024.2431042

McCann, T. V., & Lubman, D. I. (2018). Adaptive coping strategies of affected family members of a relative with substance misuse: A qualitative study. Journal of Advanced Nursing, 74(1), 100–109. 10.1111/jan.13405

O’Connor, D. (2021). Mothering through a child’s addiction journey: Linking lived experience to the lenses that shape intervention. Journal of Social Work, 21(6), 1339–1359. 10.1177/1468017320954349

Orford, J., Copello, A., Velleman, R., & Templeton, L. (2010). Family members affected by a close relative’s addiction: The stress-strain-coping-support model. Drugs: Education, Prevention and Policy, 17(Sup1), 36–43. 10.3109/09687637.2010.514801

Orford, J., Templeton, L., Velleman, R., & Copello, A. (2010). Methods of assessment for affected family members. Drugs: Education, Prevention and Policy, 17(Sup1), 75–85. 10.3109/09687637.2010.514783

Orford, J., Velleman, R., Copello, A., Templeton, L., & Ibanga, A. (2010). The experiences of affected family members: A summary of two decades of qualitative research. Drugs: Education, Prevention and Policy, 17(Sup1), 44–62. 10.3109/09687637.2010.514192

Orford, J., Velleman, R., Natera, G., Templeton, L., & Copello, A. (2013). Addiction in the family is a major but neglected contributor to the global burden of adult ill-health. Social Science & Medicine, 78, 70–77. 10.1016/j.socscimed.2012.11.036

Papamihali, K., Yoon, M., Graham, B., Karamouzian, M., Slaunwhite, A. K., Tsang, V., Young, S., & Buxton, J. A. (2020). Convenience and comfort: reasons reported for using drugs alone among clients of harm reduction sites in British Columbia, Canada. Harm Reduction Journal. 10.1186/s12954-020-00436-6

Rotunda, R. J., & Doman, K. (2001). Partner Enabling of Substance Use Disorders: Critical Review and Future Directions. The American Journal of Family Therapy, 29(4), 257–270. 10.1080/01926180126496

Williams, A. V, Marsden, J., & Strang, J. (2014). Training family members to manage heroin overdose and administer naloxone: randomized trial of effects on knowledge and attitudes. In Addiction (Abingdon, England) (Vol. 109, Issue 2, pp. 250–259). Blackwell Publishing Ltd. 10.1111/add.12360

Wilson, E. (2019). Community-Based Participatory Action Research. In Handbook of Research Methods in Health Social Sciences (pp. 285–298). Springer Singapore. 10.1007/978-981-10-5251-4_87

